# Deciphering the state of immune silence in fatal COVID-19 patients

**DOI:** 10.1101/2020.08.10.20170894

**Authors:** Pierre Bost, Francesco De Sanctis, Stefania Canè, Stefano Ugel, Katia Donadello, Monica Castellucci, David Eyal, Alessandra Fiore, Cristina Anselmi, Roza Maria Barouni, Rosalinda Trovato, Simone Caligola, Alessia Lamolinara, Manuela Iezzi, Federica Facciotti, Anna Rita Mazzariol, Davide Gibellini, Pasquale De Nardo, Evelina Tacconelli, Leonardo Gottin, Enrico Polati, Benno Schwikowski, Ido Amit, Vincenzo Bronte

**Affiliations:** Department of Immunology, Weizmann Institute of Science, Rehovot, Israel; Systems Biology Group, Department of Computational Biology and USR 3756, Institut Pasteur and CNRS, Paris 75015, France; Sorbonne Universite, Complexite du vivant, Paris 75005, France; Immunology Section, Department of Medicine, University and Hospital Trust of Verona, Verona 37134, Italy; Intensive Care Unit, Department of Surgery, Dentistry, Maternity and Infant, University and Hospital Trust of Verona, Verona 37134, Italy; The Center for Technological Platforms, University of Verona, Verona 37134, Italy; CAST- Center for Advanced Studies and Technology, University of G. D’Annunzio of Chieti-Pescara, Chieti 66100, Italy; Department of Experimental Oncology, IEO European Institute of Oncology IRCCS, Milan 20139, Italy; Microbiology Unit, Department of Diagnostics and Public Health, University and Hospital Trust of Verona, Verona 37134, Italy; Division of Infectious Diseases, Department of Diagnostics and Public Health, University and Hospital Trust of Verona, Verona 37134, Italy

## Abstract

Since the beginning of the SARS-CoV-2 pandemic, COVID-19 has appeared as a unique disease with unconventional tissue and systemic immune features. While COVID-19 severe forms share clinical and laboratory aspects with various pathologies such as hemophagocytic lymphohistiocytosis, sepsis or cytokine release syndrome, their exact nature remains unknown. This is severely impeding the ability to treat patients facing severe stages of the disease. To this aim, we performed an in-depth, single-cell RNA-seq analysis of more than 150.000 immune cells isolated from matched blood samples and broncho-alveolar lavage fluids of COVID-19 patients and healthy controls, and integrated it with clinical, immunological and functional *ex vivo* data. We unveiled an immune signature of disease severity that correlated with the accumulation of naïve lymphoid cells in the lung and an expansion and activation of myeloid cells in the periphery. Moreover, we demonstrated that myeloid-driven immune suppression is a hallmark of COVID-19 evolution and arginase 1 expression is significantly associated with monocyte immune regulatory features. Noteworthy, we found monocyte and neutrophil immune suppression loss associated with fatal clinical outcome in severe patients. Additionally, our analysis discovered that the strongest association of the patients clinical outcome and immune phenotype is the lung T cell response. We found that patients with a robust CXCR6+ effector memory T cell response have better outcomes. This result is line with the rs11385942 COVID-19 risk allel, which is in proximity to the CXCR6 gene and suggest effector memory T cell are a primary feature in COVID-19 patients. By systemically quantifying the viral landscape in the lung of severe patients, we indeed identified Herpes-Simplex-Virus 1 (HSV-1) as a potential opportunistic virus in COVID-19 patients. Lastly, we observed an unexpectedly high SARS-CoV-2 viral load in an immuno-compromised patient, allowing us to study the SARS-CoV-2 *in-vivo* life cycle. The development of myeloid dysfunctions and the impairment of lymphoid arm establish a condition of immune paralysis that supports secondary bacteria and virus infection and can progress to “immune silence” in patients facing death.

## Main

Severe acute respiratory syndrome coronavirus 2 (SARS-CoV-2) is the etiological agent of the novel coronavirus disease (COVID-19) outbreak in Wuhan city (China) [1] that is currently threatening worldwide health. Italy was the first European nation to be severely affected by COVID-19: the first death was reported on 21 of February, 2020 [2]; and as of 21^st^ July 2020 more than 35,000 and 607,781 COVID-19-related deaths were registered in Italy and worldwide, respectively [3].

Many studies highlight different, stepwise patterns of diseases progression, characterized by mild to moderate features in most of the patients, with some of them who unfortunately progress to a more severe disease stage, which can lead to acute respiratory distress syndrome (ARDS), respiratory failure and eventually death [4, 5]. The contribution of host immune system in establishing the worse prognosis has been already confirmed by several clinical observations on SARS-CoV-2 and other SARSs-dependent diseases. Indeed, lymphopenia and release of pro-inflammatory cytokines such as CXCL10 (IP10), interleukin (IL)6, IL8, IL10, tumor necrosis factor (TNF)α and C-C motif chemokine ligand (CCL)2 are enlisted as hallmark of severe SARS-CoV2 infection and correlate with adverse clinical outcome [5-7]. Accordingly, among clinical parameters associated to critical outcome, multicenter analysis on hospitalized COVID-19 patients, established among clinical parameters associated to critical outcome not only age, co-morbidities and pre-existing diseases but also immune alterations such as an increased neutrophil to lymphocyte ratio [8], hinting that pathogenic disease characteristics of the disease are worsened in sub-optimally efficient and immune dysfunctional patients.

Whether a dysregulated host immune system characterized by the coexistence between pro-inflammatory and anti-inflammatory mediators represents a key feature of COVID-19 severe progression, a clear frame of the molecular mechanisms driving this imbalance is not elucidated yet. Indeed, ARDS experienced by COVID-19 patients has a unique signature that differs from ARDS caused by any other infective or traumatic insults [9]. More specifically, the increase in cytokine release in peripheral blood, often associated with disease severity [10] and commonly defined as “cytokine storm”, is only partially involved in COVID-19 patients. Indeed, IL6 plasma levels in COVID-19 severe patients are 10 to 40 fold lower than previously reported ARDS patients, and 1,000 fold lower compared to patients facing cytokine release syndrome following treatment with chimeric antigen receptor T cells [11, 12]. Thus, it is conceivable that SARS-CoV-2 infection may hijack host immune system in order to impair anti-viral immunity and trigger a chronic inflammation characterized, but not limited, by the accumulation of peculiar inflammatory cytokines that participate in acute lung injury in severe COVID-19 patients. Recent literature explored the ability of CoVs to skew cytokine release by affecting IFN-dependent, anti-viral response towards other inflammatory pathways sustaining the activation of inflammasome [13]. Noteworthy, delayed IFN-I signaling impairs antigen-specific T cell responses and promotes high cytokine secretion in lung by incoming monocytes, resulting in vascular leakage and fatal disease in SARS-CoV-infected mice [14]; furthermore, type I IFN, T cells, and signal transducer and activator of transcription 1 (STAT1) are required for virus clearance and disease resolution in a mouse model of SARS-Cov 2 infection [15] and impaired IFN I activity results in worse outcome in human COVID-19 infected patients [16].

Severe COVID-19 patients display some shared features of sepsis, including secretion of inflammatory cytokines, neutrophil hyper-activation, reduced function of natural killer (NK) and dendritic cells (DC), altered monocyte activation and lymphopenia [7, 17]. Several high dimensional phenotypic and molecular approaches were deployed in order to dissect the biology of virus-immune system interaction during COVID-19 pathogenesis [7, 16, 18-22]. These analyses were performed on peripheral blood and peripheral blood mononuclear cells (PBMCs) isolated from patients with different disease severity. This strategy streamlines sample accessibility, identification of new peripheral predictive biomarkers, and allows comparison within different studies with the caveat of not considering the local microenvironment in which the virus is acting. Taking together, all these studies highlight the presence of an IFN signature in mild to moderate patients whereas evidence a sustained emergency myelopoiesis associated to an increase in immature neutrophils and monocytes with immune suppressive features in critically ill patients. Unfortunately, none of the past studies analyzed the immune regulatory properties of myeloid cells at functional level. A specific genetic locus including immune related genes (such as C-X-C motif chemokine receptor 6 - CXCR6), was found to be associated with worse prognosis in COVID-19 patients [23]. Other studies exclusively performed on broncho-alveolar lavage fluids (BAL), elucidated the sustained interplay between macrophages releasing inflammatory cytokines and lung epithelial cells in more severe COVID-19 disease stages [21], whereas pointed out highly clonally expanded CD8^+^ T cells in moderate patients [22]. Here we define a complete atlas of COVID-19 patients’ immune landscape integrating both local (lung) and systemic (blood) tissues, harmonizing molecular (single cell RNA sequencing, scRNA-seq), functional and clinical data, in order to dissect the complex interplay established by SARS-CoV-2 with host immune system. We find in severe patients the establishment of innate and adaptive dysfunctions, including loss of immune-suppression by various blood myeloid cells and the replacement of lung memory CD8^+^ T cells by naive T cells, suggesting a state of “immune silence” that correlates with a severe clinical manifestation and fatal outcome.

### Establishment of BAL and blood-derived immune cell atlas obtained from COVID-19 patients

To gain insights into the immune deviation induced by SARS-CoV-2 virus in COVID-19 patients, we performed scRNA-seq analysis on BAL and matched peripheral blood samples obtained from 21 severe, COVID-19 patients admitted to Intensive Care Units (ICU) and on peripheral blood of 6 mild SARS-CoV-2 positive patients and 5 healthy donors (Figure 1a). Immunological features were assessed on the same cohorts, integrating 4 more mild SARS-CoV-2 patients, by multiplex ELISA, multiparametric flow cytometry and functional assay (see Methods). All the patients were hospitalized at the University Hospital Integrated Trust of Verona. The clinical characteristics of enrolled patients and healthy controls are summarized in table 1.

**Figure 1.**
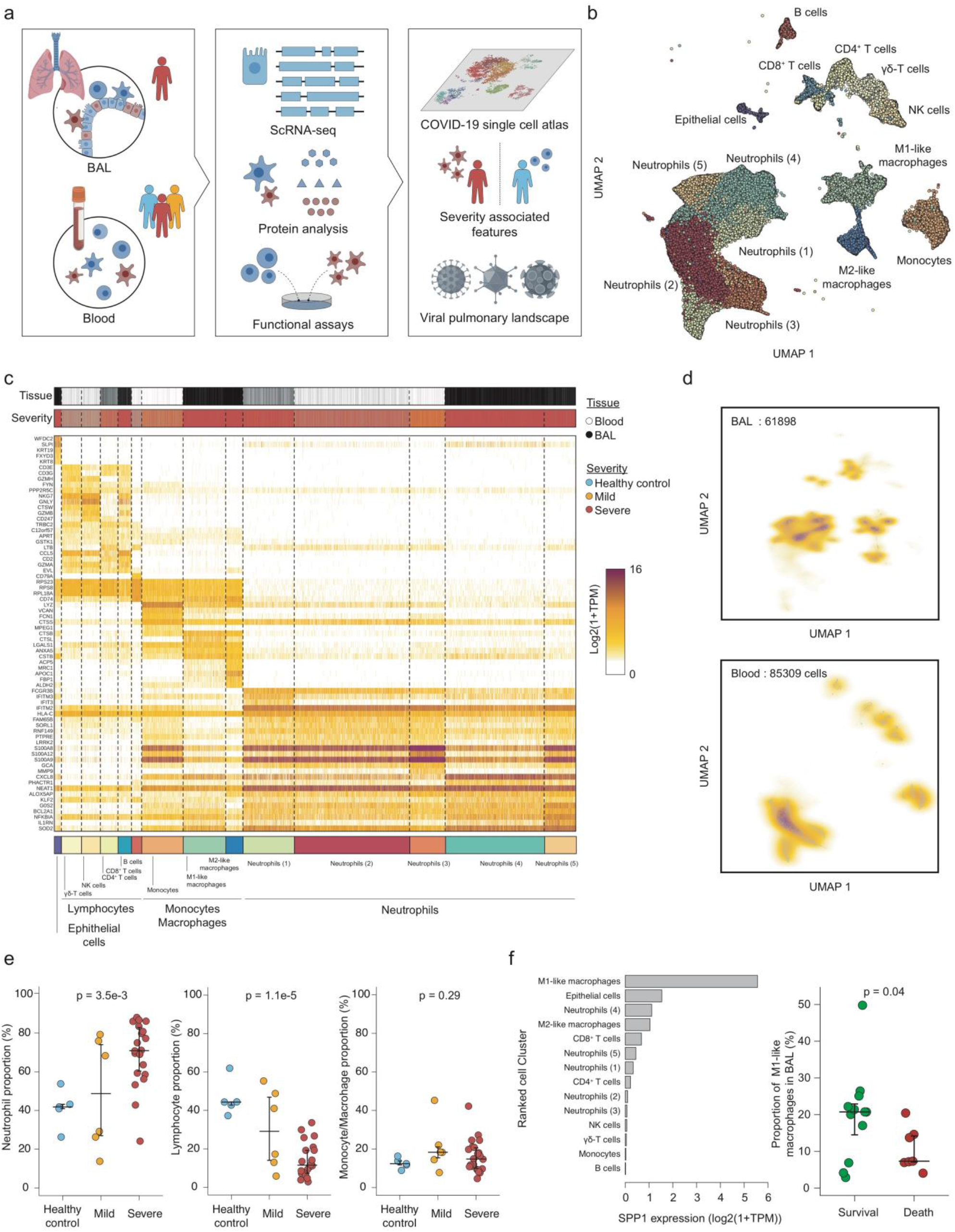
Establishment of BAL and blood-derived immune cell atlas obtained from COVID-19 patients. **(a)** Description of the cohort. **(b)** Two-dimensional UMAP embedding of the scRNA-seq data. Dots (cells) are colored according to their respective metacluster (Epithelial cells, lymphocytes, neutrophils and monocytes/macrophages). **(c)** Expression heatmap of the 14 significant clusters detected in our scRNA-seq dataset. The top 5 best markers were selected for each cluster. **(d)** Two-dimensional density plot of the UMAP embedding of the BAL (upper panel) or the blood (lower panel) cells. **(e)** Proportion of neutrophils, lymphocytes and monocytes/macrophages in blood samples across patient status. For each of the cell types, an ANOVA test was performed and corrected for multiple-testing by Bonferroni correction. Median and 5%-95% theoretical quantiles are shown. **(f)** Expression of the SPP1 (osteopontin) gene across cell types (left panel) and distribution of M1 macrophages among total BAL cells based on severe patient clinical outcome (right panel). A two-sided Welch’s t-test was performed to compare proportion between the two groups of patients (t = 2.2 with a degree of freedom equal to 16.5). Median and 5%-95% theoretical quantiles are shown.

**Table 1:** Clinical characteristic of enrolled patients and healthy controls. IQR denotes interquartile range, PCO2 carbon dioxide partial pressure, PO2 oxygen partial pressure, FiO2 fraction of inspired oxygen, P/F PO2/FiO2, CFU denotes colony forming units.

| **CHARACTERISTICS** | **Healthy Controls**<br>**N=5** | **Mild Patients**<br>**N=10** | **Severe Patients (ICU)**<br>**N=21** |
| --- | --- | --- | --- |
| **Anagraphic** |  |  |  |
| Age, yr: Median (IQR) | 66 (64-73) | 69 (56-80) | 67 (58-70) |
| Male, no. (%) | 4 (80) | 6 (60) | 17 (81) |
| **Coexisting disorder, no. (%)** |  |  |  |
| Any | 2 (40) | 10 (100) | 17 (81) |
| Obesity | 0 (0) | 2 (22) | 3 (14) |
| Hypertension | 2 (40) | 10 (100) | 11 (52) |
| Diabetes | 0 (0) | 3 (30) | 7 (33) |
| Chronic obstructive pulmonary disease | 0 (0) | 2 (20) | 1 (5) |
| Cardiovascular disease | 0 (0) | 5 (50) | 3 (14) |
| Cronic kidney disease | 0 (0) | 2 (20) | 1 (5) |
| Active malignancies | 0 (0) | 0 (0) | 2 (10) |
| **Median (IQR) interval from syntoms onset (S.O.) and Outcome** |  |  |  |
| Days from S.O. to Hospitalization | - | 5 (2-6) | 6 (4-7) |
| Days from S.O. to ICU admission | - | - | 7 (6-10) |
| Days from S.O. to dismissal from ICU | - | - | 38 (23-45) |
| Days from S.O. to dismissal from Verona Hospital (alive or dead) | - | 21 (13-24) | 43 (32-66) |
| Outcome, no. deaths (%) | - | 1 (10) | 8 (38) |
| **Clinical features at sampling** |  |  |  |
| APACHE score, Median (IQR) | - | - | 23.5 (15-28.5) |
| SOFA score, Median (IQR) | - | 2 (0.8-3.3) | 6 (4-7) |
| pCO2 [35-40 mmHg], Median (IQR) | - | 34 (31-41) | 48 (41-52) |
| pO<sub>2</sub> [80-100 mmHg], Median (IQR) | - | 59 (55-64) | 77 (69-97) |
| FiO<sub>2</sub> %, Median (IQR) | - | 26 (21-31) | 60 (50-90) |
| P/F ratio mmHg, Median (IQR) | - | 213 (194-267) | 146 (71-177) |
| **Laboratory findings at sampling, Median (IQR)** |  |  |  |
| Hemoglobin (135-160 g/L) | 151 (145-153) | 120 (98-137) | 104 (89-112) |
| Leukocytes (4,3-10 10<sup>9</sup>/L) | 6.7 (6.2-8.8) | 5.6 (5.2-6.4) | 12.8 (9.6-15.8) |
| Neutrophils (1,8-8 10<sup>9</sup>/L) | 3.8 (3.4-4.9) | 3.8 (3.1-4.6) | 10.3 (7.7-13.4) |
| Lymphocytes (1,2-4 10<sup>9</sup>/L) | 2.2 (1.7-2.8) | 1.3 (1-1.8) | 1.2 (0.7-1.4) |
| Monocytes (0,2-1 10<sup>9</sup>/L) | 0.7 (0.4-0.8) | 0.5 (0.4-0.8) | 0.7 (0.4-1.1) |
| Eosinophils (<0,45 10<sup>9</sup>/L) | 0.3 (0.3-0.4) | 0.1 (0-0.1) | 0.2 (0.1-0.4) |
| Basophils (<0,2 10<sup>9</sup>/L) | 0.05 (0.04-0.08) | 0.02 (0.01-0.04) | 0.04 (0.02-0.07) |
| Platelets (150-400 10<sup>9</sup>/L) | 233 (216-234) | 248 (160-311) | 285 (217-368) |
| C-reactive protein [inf. a 5 mg/L] | - | 24 (22-56) | 131 (75-158) |
| P-Ferritin [30 - 300 µg/L] | - | 947 (342-1368) | 1262 (735-1647) |
| P-D-Dimer [inf. a 500µg/L] | - | 1600 (690-2501) | 2250 (1319-3684) |
| Trombin clotting time (pt) 0,82-1,17 INR | - | 1.06 (0.98-1.1) | 1.14 (1.09-1.25) |
| P-fibrinogen [2,00 - 4,00 g/L] | - | 4.2 (3.3-5) | 6.8 (5.4-7.8) |
| **Microbiology analysis on BAL at sampling** |  |  |  |
| Lung infection (CFU>10<sup>4</sup>), no. (%) | - | - | 10 (48) |
| Pseudomonas lung infection (CFU>10<sup>4</sup>), no. (%) | - | - | 7 (33) |

IQR denotes interquartile range, PCO<sub>2</sub> carbon dioxide partial pressure, PO<sub>2</sub> oxygen partial pressure, FiO<sub>2</sub> fraction of inspired oxygen, P/F PO<sub>2</sub>/FiO<sub>2</sub>, CFU denotes colony forming units.

Following strict quality controls (Figure S1a-c), cells were analyzed using the Pagoda2 pipeline [24] and clustered using Leiden community detection method [25]. The number of analyzed high quality cells was comparable between groups (Figure S1d). Fourteen significant cell clusters (more than 1% of the cells) were identified, and gathered into four major cellular subsets based on their mean expression profiles (Figure 1b and C, Figure S1e): epithelial cells, lymphocytes, monocytes/macrophages and neutrophils. The epithelial cell compartment contained only one cell cluster, which was characterized by the expression of WFDC2, SPLI and keratin genes, like KRT8 and KRT19. 5. Different lymphocyte clusters were identified, namely B cells (CD79A, CD74), NK cells (CD247, GZMB, GNLY), CD8^+^ T cells (GZMA, CD8A), CD4^+^ T cells (LTB) and gamma-delta (γδ)-T cells (GZMH). Only three clusters of the monocyte/macrophage compartment were depicted in our dataset, monocytes (LYZ, VCAN, FCN1), M1-like (CTSB, CTSL) and M2-like macrophages (MRC1, ACP5, FBP1). Lastly, significant diversity of the neutrophil compartment was observed with 5 uncovered clusters. Those clusters could be differentiated based on their expression of key markers such as CD16B (FCGR3B) mostly expressed in neutrophil clusters (1), (2) and (3), interferon response genes (IFITM3 and IFIT3) in cluster (1), S100 calcium binding proteins (S100A8, S100A9 and S100A12) in clusters (3) and (5), CXCL8 in cluster (4) and inflammatory response genes (NFKBIA, IL1RN and SOD2) in cluster (5).

We observed a strong tissue specificity in the cell clusters distribution (Figure 1c and d). For instance, neutrophil clusters (4) and (5) were BAL specific, while clusters (2) and (3) were blood specific and cluster (1) could be found in both tissues. As expected, M1-like and M2-like macrophages could only be identified in BAL samples while monocytes were blood specific. Epithelial cells were limited to the BAL samples with low representation of the total cell population (1.1% of total cells).

We next looked for possible associations between the proportion of cell clusters and the clinical status of the patient. We observed that severe patients exhibited a significantly higher proportion of neutrophils in their blood samples compared to mild patients and healthy controls (Figure 1e, left panel). In contrast, the lymphocyte proportion was decreased in severe patients compared to mild patients and healthy controls (Figure 1e, middle panel) while the monocyte/macrophage compartment was not affected by disease severity (Figure 1e, right panel). As scRNA-seq is prone to biases for population proportion estimation, we validated our findings by performing blood cell counting and systemically looking for differences between the 3 groups of patients. Consistently with our scRNA-seq analysis, we observed significant differences in the neutrophil and lymphocyte population, but also a decreased erythrocyte number in severe patients (Figure S1f and g). No other cell population was significantly affected by the disease (Figure S1f).

COVID-19 is characterized by an excessive inflammatory response that is sometimes referred to “cytokine storm” and is deemed to drive the disease pathogenesis. As inflammatory macrophages are suspected to be the main producer of inflammatory cytokines such as IL6 and IL1β, we looked for possible enrichment of M1 macrophages in the BAL from patients who died due to COVID-19. While this cluster was indeed a significant producer of inflammatory cytokines such as osteopontin (SSP1, Figure 1f left panel), it was surprisingly associated with the survival of the patients (Figure 1f right panel), suggesting it as a predictive biomarker for better prognosis. Lastly, we investigated the concentration of cytokines in the plasma and systemically looked for differential concentration between the classes of patients. Following multiple testing correction, we observed that only three cytokines were significantly affected by the patient status: Vascular Endothelium Growth Factor - alpha (VEGF-A), IL6 and Interleukin -1 receptor Antagonist (IL1RA, encoded by the IL1RN gene). All three exhibited higher concentrations in severe and mild patients’ plasma compared to healthy control, whose levels were close to detection limit (Figure S1h to i).

Altogether, we comprehensively profiled more than 150.000 immune cells from blood and BAL sampled from COVID-19 patients. Coarse-grained clustering allowed us to detect a severity associated neutrophilia and lymphopenia, but also a SPP1^+^ M1-like macrophage population associated with severe patients’ survival. Interestingly, only the concentration of these three cytokines was associated with disease severity, suggesting that massive cytokine release in blood is not present in COVID-19 patients. However, the restricted size of our cohort significantly limits the statistical power of our analysis and might hinder the detection of disease-associated variables.

### Resting neutrophils are replaced by multiple SARS-Cov2 associated neutrophil types

Neutrophils are the most common white blood cells and are the first cells to migrate to the site of infection. Our dataset contains 42.238 high quality blood neutrophils, therefore allowing an in-depth analysis. We performed a refined clustering of neutrophils, which identified 10 different clusters (Figure 2a and b), including a strongly distinct and rare subtype of CD66b (CEACAM8) and antibacterial peptide (LTF, DEFA3) expressing neutrophils, probably corresponding to low density neutrophils (LDNs) and were therefore defined as LDN-like cells. Among the other clusters, we observed both a resting neutrophil cluster (ICAM1, CXCL8) and an array of activated neutrophil clusters. Among them we identified an Interferon Stimulated Genes (ISGs; RSAD2, OAS2, IFIT1), a serine protease inhibitor (PI3 and SLPI), and a chemokine (CCL4, CCL3L3) expressing clusters, suggesting a strong heterogeneity of the neutrophil polarization across patients.

**Figure 2.**
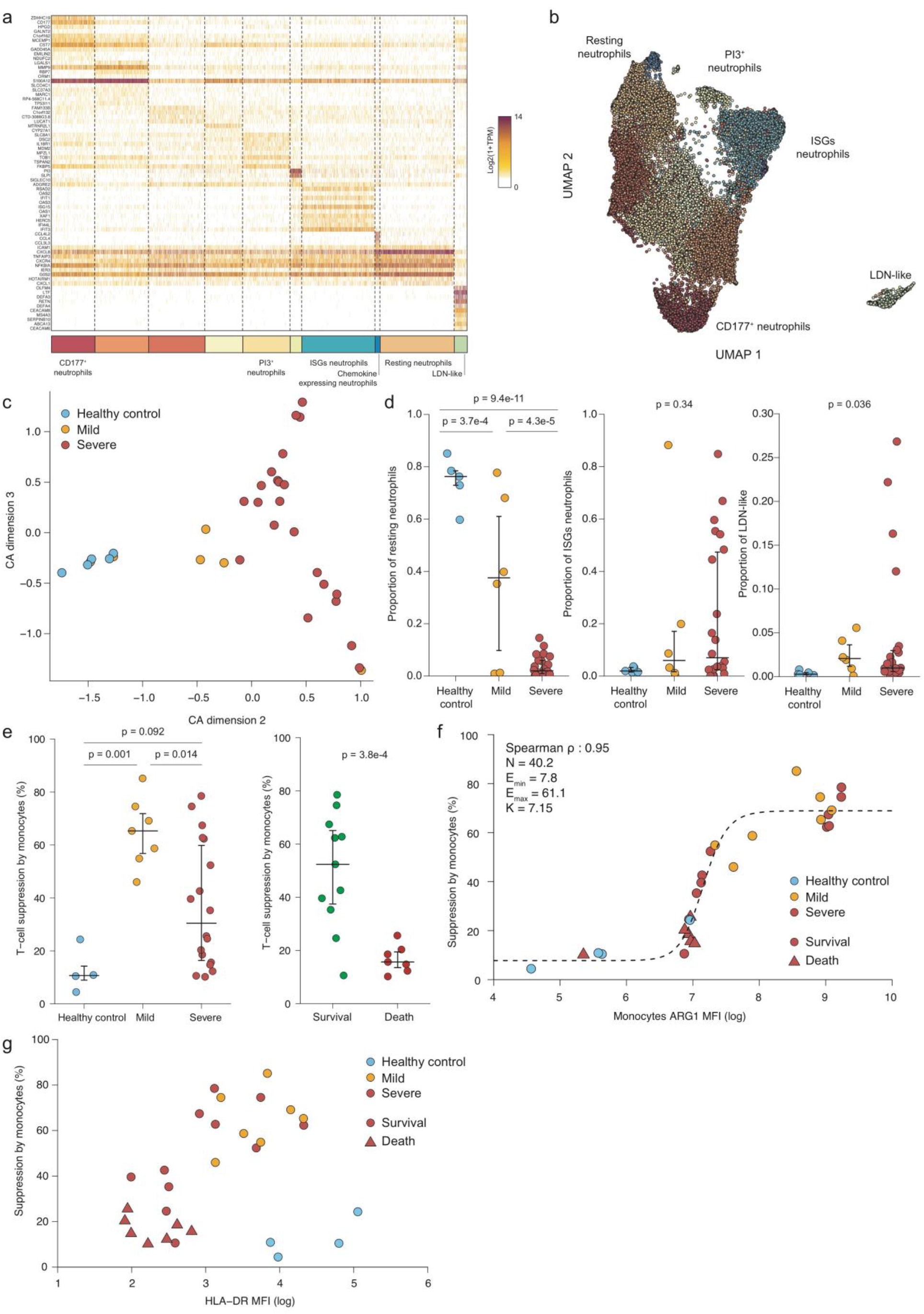
ScRNA-seq and functional analysis of blood myeloid cells reveal unique features associated to patient status and outcome. **(a)** Expression heatmap of the 10 clusters identified among the blood neutrophils. **(b)** Two-dimensional UMAP embedding of the blood neutrophil. Cells are colored according to their cluster. **(c)** Scatter plot of the Correspondence Analysis of the blood neutrophil populations. **(d)** Proportion of resting neutrophils (left panel), ISGs neutrophils (middle panel) and LDN-like (right panel) among blood neutrophils according to patient clinical status. A Tukey’s range test was used in the left panel while Kruskall-Wallis rank test was used in both middle and right panel. Median and 5%-95% theoretical quantiles are shown. **(e)** T-cell suppression ability of CD14+ monocytes according to clinical status (right) or to ICU outcome (severe patients only, right panel). A Tukey’s range test was used in the left panel while a two-sided Welch’s t-test was performed in the right panel (t = 4.9 with a degree of freedom equal to 11.8). Median and 5%-95% theoretical quantiles are shown. **(f)** Association between CD14+ monocytes ARG1 Mean Fluorescence Intensity (MFI) and immune suppression. The dashed line corresponds to the fitted Hill-like function. **(g)** Association between CD14+ monocytes HLA-DR Mean Fluorescence Intensity (MFI) and immune suppression.

To identify any trend in the neutrophil compartment composition in an un-supervised fashion, we used the correspondence analysis (CA), a method similar to Principal Component Analysis but adapted to categorical data (see Methods). CA second component was able to stratify healthy controls, mild and severe patients (Figure 2c, S2a and S2b). By computing the correlation between CA dimension 2 and the proportion of each neutrophil cluster we observed that severe, and to a lesser extent mild patients, were associated to a replacement of resting neutrophils by multiple clusters including the ISGs, CD177 and PI3 expressing neutrophils (Figure 2d, S2c). Interestingly, LDN-like cells were only detected in both mild and severe patients, albeit at a low level (less than 5% of neutrophils) except in four severe patients (Figure 2D right panel). We systemically computed Pearson’s correlation between CA dimension 2 and each measured biological and clinical variable and identified IL6 and IL1RA concentration as the most positively correlated variables, with erythrocyte and partial CO2 concentration (pCO2) negatively correlating with CA dimension 2 (Figure S2d). Altogether, our refined analysis of blood neutrophils revealed that resting neutrophils are replaced by various neutrophil clusters in both mild and severe patients.

### Functional analysis of blood myeloid compartment reveals that immuno-suppression is a major predictor of COVID-19 patients’ survival

Blood neutrophil compartment is widely affected in COVID-19, as highlighted by our scRNA-seq analysis. However, these cells are naturally over-represented in blood and most severe COVID-19 patients suffer from neutrophilia, therefore limiting the number of cells other than neutrophils sequenced in our blood samples and our ability to study them by scRNA-seq. Recent evidence suggested the presence of monocyte alteration in SARS-CoV-2 infected patients, mostly associated with the expansion and accumulation of immunosuppressive monocytes [26]. Due to the low number of blood monocytes sequenced in our dataset (9103 cells, corresponding to less than 300 cells per patient) and the limited ability of scRNA-seq to provide functional information, we purified circulating CD14^+^ monocytes from fresh blood of some study subjects and used them to perform T-cell immuno-suppression assays. Both cellular and supernatant-associated immune-suppressions were assessed. In addition, immuno-suppressive activities of both normal density neutrophils (NDN) and LDN supernatants were also measured.

All samples caused some degree of T cell suppression, with monocytes and monocyte supernatants exhibiting similar activity, while LDN and NDN exhibited the highest and lowest suppression activity, respectively (Figure S2e). Suppression rate by monocytes was significantly lower in healthy controls compared to both mild and severe patients but severe patients displayed a higher variance than mild patients, with the suppression rate ranging from 10% to nearly 80% (Figure 2e left panel). Surprisingly, this heterogeneity could be partly explained by the clinical outcome of the severe patients: while severe patients who survived displayed a high T-cell immuno-suppression by monocytes, monocytes of deceased patients were unable to dampen T-cell proliferation (Figure 2e, right panel). This inverse association between immuno-suppression and patient survival was not limited to monocytes as it could be observed, albeit less significantly, with both monocyte and LDN supernatants (Figure S2f).

Myeloid cells suppress T-cell activation through multiple strategies including anti-inflammatory cytokine secretion, nutrient depletion or immune checkpoint engagement [27]. To gain further insight in COVID-19 immune landscape, we profiled monocyte expression of PD-L1, ARG1 and HLA-DR by flow cytometry (Figure S2g). We observed a clear relation between mean ARG1 expression by monocytes and monocyte immunosuppressive function (Spearman ρ = 0.95; Figure 2f), which could be fitted using a modified Hill function (see Methods), revealing an extremely strong Hill coefficient (n=40.2). HLA-DR expression was also associated to immune suppression, but in a different manner compared to ARG1 (Figure 2g). HLA-DR mean expression and immune suppressive activity clustered three groups of patients: healthy controls with a high HLA-DR expression and low immuno-suppression; mild patients and severe patients who survived with both high suppression and high HLA-DR expression; a third group of severe patients with low suppression and HLA-DR expression. As more than half (7/12) of the patients from the last groups died, we hypothesize that this cluster corresponds to patients suffering from terminal immune dysfunctions and therefore at higher risk of fatal outcome. Lastly, we observed a limited association between PD-L1 and immuno-suppression (Spearman ρ= 0.57) (Figure S2h). Furthermore, the concentration of 20 different cytokines, including both pro-inflammatory (IL6, TNFα) and anti-inflammatory ones (IL10) was assessed in monocyte supernatant; however, no cytokine highly correlated with immune suppression (absolute Spearman ρ lower than 0.4; Figure S2i). In summary, the immuno-suppressive activity of monocytes and other blood myeloid cells is a strong predictor of severe patient survival and is primarily associated with ARG1 expression, and to a lesser extent with PD-L1 but not with any specific cytokine secretion.

### COVID-19 affects blood and lung lymphocyte compartments in a severity-dependent manner

The lymphocyte compartment is extremely heterogeneous and dynamic, since it contains various cell types with properties and functions that can evolve upon inflammation and infection. By re-clustering cells identified as lymphocytes, we were able to obtain a finer picture of their heterogeneity. We identified 14 clusters, including several effector and memory T cells, naïve T cells and activated γδ-T cells (Figure 3a and S3a). Interestingly we were able to identify a cluster of B cells expressing high level of TCF4 that was specific to the blood of patient 8, a patient suffering from Chronic Lymphocytic Leukemia (CLL). We assumed that those cells were neoplastic and were thus removed from the analysis. As expected, significant differences could be observed between blood and BAL lymphocytes, with memory, effector and dividing T cells mostly found in the BAL and NK cells in the blood (Figure 3a, right panel).

**Figure 3.**
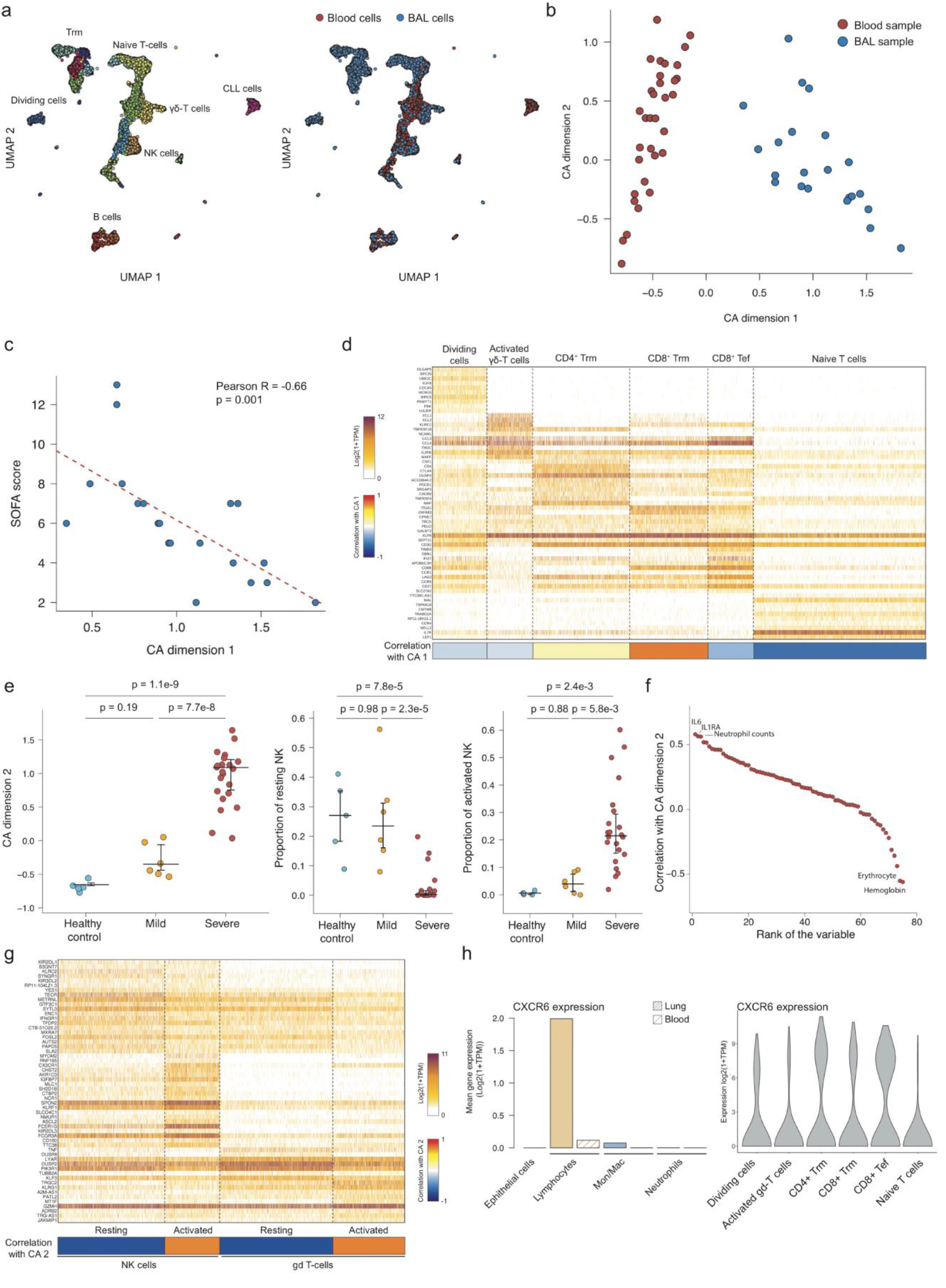
ScRNA-seq analysis of blood and BAL lymphocytes identify cellular. **(a)** Two-dimensional UMAP embedding of lymphocytes colored according to their cluster (left panel) or their tissue (right panel). **(b)** Correspondence Analysis of blood and BAL lymphocytes**. (c)** Association between SOFA score and CA dimension 1 score of BAL samples. **(d)** Expression heatmap of the BAL lymphocytes belonging to the cell clusters that are associated with CA dimension 1. The 10 best markers are shown for each cluster. **(e)** Distribution of CA dimension 2 score of blood samples according to clinical status (left panel), and proportion of resting (middle panel) and activated NK (right panel) according to clinical status. Tukey’s range test was used to compute the shown p-values. Median and 5%-95% theoretical quantiles are shown. **(f)** Ranked Pearson correlation between biological features and CA dimension 2. **(g)** Expression heatmap of the blood lymphocytes belonging to the cell clusters that are associated with CA dimension 2. The 20 best markers are shown for each cluster. **(h)** Mean expression of CXCR6 across different tissues and cell types (left panel) and among the different BAL lymphocytes clusters associated with CA dimension 1.

To identify trends in the lymphoid compartment composition in an un-supervised manner, we used the CA, as described above. The first CA dimension of the lymphoid population perfectly separated the blood and BAL samples (Figure 3b and S3b) and seemed to capture a major trend in the BAL lymphocyte population. We therefore computed the correlation between this dimension and the various clinical features measured and noticed a striking negative correlation with the Sequential Organ Failure Assessment (SOFA) score (Figure 3c) but not with other variables (Figure S3c). In opposition, a parallel analysis performed on the blood of severe patients identified no clinical parameters associated with CA dimension 2 (Figure S3d), strengthening the importance to analyze the main district affected by the disease. Among the six major BAL lymphocytes clusters (>5% of total BAL lymphoid cells clusters) CD8+ T resident memory (CD8^+^ Trm, expressing ZNF683 and ITGA1) had the strongest positive correlation with CA dimension 1 (R=0.79), followed by CD4^+^ T resident memory (CD4^+^ Trm) (R=0.17). Interestingly, CD4^+^ Trm expressed several immune checkpoints such as CTLA4 and PD-1 (encoded by PDCD1), suggesting that they can participate to the immune suppressive landscape (Figure 3d). The naïve T-cell cluster (IL7R, LEF1) had the strongest negative correlation with the CA dimension 1, and therefore the strongest positive association with the SOFA score (R=-0.66). Three other clusters were negatively associated with CA dimension 1: effector CD8^+^ T cells (R=-0.30, LAG3 and CD27), dividing T cells (R=-0.16, MCM10, E2F8) and activated γδ-T cells (R=-0.13, XCL1/2 and TRDC).

To test whether CA was also able to capture meaningful variations in blood lymphocyte population, we analyzed potential association between patient status and CA dimension 2 as it seemed to capture an important trend in blood samples (Figure 3b). Interestingly, CA dimension 2 was significantly associated with patient clinical status with a higher value being specific for severe patients (Figure 3e, left panel). This dimension was associated with two populations of NK cells and two populations of γδ-T cells (Figure 3e and 3g), i.e. resting/activated NK cells, and resting/activated γδ-T cells. Both activated NK and γδ-T cells were associated with severe COVID-19 patients while resting cells were found in healthy controls and mild patients only (Figure 3e and S3e). Activated NK and γδ-T cells were associated with an increased expression of cytotoxicity genes and activation markers such as PRF1, NKG7, KLRG1 and CD247 (Figure 3g) while resting NK cells were featured by a higher expression of the inhibitory KIR receptors KIR2DL1 and KIR3DL2 and resting γδ-T cells by a higher expression of TNFα and DUSP8 (Figure 3g). Consistently with our initial analysis of the cytokine and blood cell count data, we found that CA dimension 2 was positively associated with IL1RA plasma concentration and neutrophil counts and negatively associated with erythrocyte count and hemoglobin concentration (Figure 3f). Taken together, our analysis of the lymphoid compartment revealed that the presence of a naïve T-cell population in the BAL is associated with high clinical severity, while the blood of severe COVID-19 is characterized by the activation of NK and γδ-T cells.

### rs11385942 may act trough the regulation of the memory lymphoid cell migration

Expression profiles generated by scRNA-seq can be overlaid with genome-wide association studies (GWASs) to pinpoint specific cell types and identify potential cellular and molecular mechanisms explaining the described genetic associations [28]. In addition to the ABO group locus, a recent study found that another genomic locus is associated with the development of severe forms of COVID-19 [23]. However, how this locus contributes to the pathology is unclear. As six different genes were covered by the peak association (SLC6A20, LZTFL1, CCR9, FYCO1, CXCR6, and XCR1), we looked at their expression in the four different cellular compartments in both blood and BAL samples. Strikingly, only CXCR6 was expressed at a detectable level (Figure 3h left panel, S3f) and specifically in BAL lymphocytes. When analyzing the expression pattern among BAL lymphocytes, it appeared to be expressed mostly by effector and memory T cells and not by the naïve T cell cluster (Figure 3h, right panel). As the risk allele is associated with a decreased expression of CXCR6 and that CXCR6 is expressed by key protective populations, we advance the hypothesis that patients with the risk allele have a lower amount of the protective T cell populations in the lung, therefore increasing the risk of developing severe COVID-19 forms.

### Viral landscape of COVID-19 patients affects their immune profile

At the time of ICU admission, most COVID-19 patients exhibited a low viral load in the lung, suggesting that the virus had been mostly eliminated and that at this time the pathology was mainly driven by an inappropriate immune response rather than by viral replication. To validate this hypothesis, we applied our recently published tool, Viral-Track [18] on the BAL sequencing data in order to quantify the SARS-CoV-2 viral load and detect possible secondary viral infections.

Quantification of SARS-CoV-2 viral reads revealed that in most of severe patients (17/21), no SARS-CoV-2 reads could be found (Figure 4a). Out of the 4 SARS-CoV-2 positive patients, three had low levels of viral reads (few hundreds to few thousands reads) while patient 8 displayed more than 300,000 SARS-CoV-2 reads. Consistently with our previous scRNA-seq study of SARS-CoV-2, we observed that most reads were located in the very 3’ end of the viral genome (Figure 4b), probably due to the 3’ bias of our scRNA-seq method and to nested replication process of the virus [29]. Surprisingly, we also found that two patients had a significant amount of Herpex Simplex Virus 1 (HSV-1) reads (NCBI reference number: NC_001806) with respectively 265.392 and 46.229 viral reads in patients 4 and 25, respectively (Figure 4c). Coverage analysis revealed dozens of peaks that corresponds to viral genes, suggesting an active transcription of the viral genes, and likely an active viral replication (Figure 4d). We validated this finding by performing a multiplex PCR test for four different Herpesviridae, HSV-1, HSV-2, Human Cytomegalovirus (HCMV) and Varicella-Zoster Virus (VZV) on samples from patients 8 (CLL patient, negative control), 13 (negative control) and patient 25 (Figure S4a). None of the viruses was detected in samples from patient 8 and 13 but HSV-1 was specifically detected in BAL samples of patient 25 that were collected at two different time points during ICU permanence.

**Figure 4.**
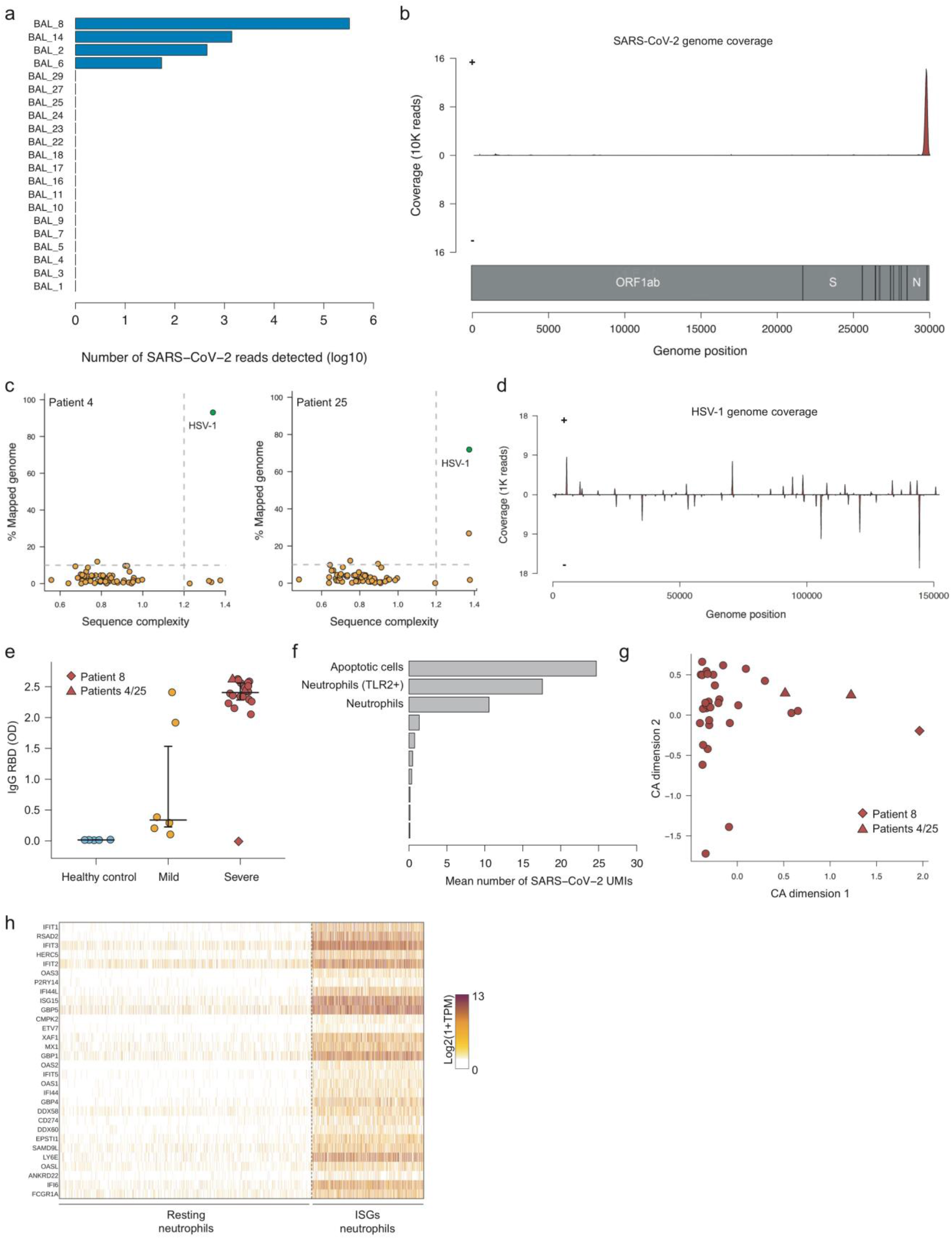
Analysis of severe COVID-19 patients viral landscape. **(a)** Number of SARS-CoV-2 UMIs in each BAL sample. **(b)** Coverage plot of SARS-CoV-2 genome. **(c)** Viral-Track analysis of patients 4 and 25. **(d)** Coverage plot of HSV-1 genome. **(e)** Quantification of IgG targeting the RBD domain of the SARS-CoV-2’s spike protein. OD: optical density. **(f)** Mean number of SARS-CoV-2 UMIs across patient 8 cell clusters. **(g)** CA analysis of total blood cell population. **(h)** Expression heatmap of cells belonging to resting or ISGs neutrophils.

We then looked for a possible explanation about the unexpectedly high SARS-CoV-2 viral load of patient 8. As mentioned before, it appeared that patient 8 suffered from CLL, a B cell malignancy characterized by the accumulation of small, mature-appearing lymphocytes in the blood, the bone marrow and in the lymphatic systems [30]. CLL patients often suffer from immune deficiency, and more precisely, from hypogammaglobulinaemia, i.e a reduction in maturated and high-affinity antibodies. We therefore quantified the amount of IgG produced toward the RBD region of the viral spike protein (Figure 4E): while healthy controls and most of the mild patients lacked any RBD-targeting IgG, all severe patients but patient 8 had high levels of these immunoglobulins. Therefore, the lack of an efficient antibody response might have prevented the complete clearance of the virus from the lung in this patient. We tried to look which BAL cells from patient 8 were infected by SARS-CoV-2. We observed that a reduced number of cells exhibit an extremely high number of viral UMIs (several hundreds to several thousands), but interestingly those cells were mostly apoptotic cells (high levels of mitochondrial reads), neutrophils or in some rare case, cells that expressed extremely low amount of human reads (Figure 4f, S4b-c). Lastly, we looked for the possible effects of high-viral loads on the immune system state: for this purpose, we performed a correspondence analysis on all blood cell populations. We observed that the first dimension was associated with the high viral load, as patients 4 and 8 had the highest score among all patients. Furthermore, this CA dimension correlated with a subset of blood neutrophils that specifically expressed Interferon Stimulated Genes (ISGs) such as IFIT1, RSAD2 and OAS3 (Figure 4h) but also PD-L1 (CD274), suggesting that a high viral load in the lung can significantly influence the blood immune landscape in COVID-19 patients.

## Discussion

The real boundaries between sepsis induced by either SARS-CoV-2 or bacteria are still ill defined and need to be further demarcated. Indeed, hyper-inflammation associated with immune suppressive state are clinical characteristics shared among these pathologic conditions, which underscore a susceptibility state defined as sepsis-induced immunoparalysis [31]. In this disease phase, innate and adaptive dysfunctions cooperate for the ineffective clearance of the pathogen, vulnerability to secondary infections and reactivation of latent viruses [32].

Immunoparalysis is likely the ground for the HSV-1 superinfection observed in some ICU patients in our study. Recently, scRNA-seq unveiled a cluster of immature CD14^+^ monocytes, with low HLA-DR and expressing MPO, PLAC8 and IL1R2, in the blood of COVID- 19 patients and similar cells were reported in sepsis [20, 33]. These findings shed light on the practice to measure the levels of HLA-DR in monocytes as marker for sepsis, preconized but never definitively proven as mortality biomarker for severe sepsis, especially in ICU patients [34, 35]. Considering the emerging cell heterogeneity, HLA-DR levels are insufficient to reach a predictive value for patient mortality, if not associated with functional analyses. Indeed, our study shows that HLA-DR reduction in monocytes, hallmark of many COVID-19 patients, can predict patient outcome only when is associated with an impairment of their immune regulatory properties (Figure 2g). More importantly, we found that loss of function in myeloid cells mirrors an immune pathological status progressing from immune paralysis to “immune silence” associated with higher susceptibility to death event (Figure 2e, Figure S2f). CD14^+^HLA-DR^low^ monocytes and ARG1^+^MPO^+^BPI^+^, low density pre-neutrophils are expanded in severe COVID-19 patients, a likely consequence of the pervasive emergency myelopoiesis triggered by SARS-Cov2 infection [33]. Based on gene expression profiles, these cell subsets were deemed to have immune suppressive features, which were not functionally addressed in the study. To the best of our knowledge, we show for the first time that immune suppression is a hallmark of COVID-19 evolution (Figure 2e) and could be the basis for gradual loss of effector/memory in favor of naïve T-cells (Figure 3d). Moreover, we defined a correlation between ARG1 presence and the immunosuppressive activity of monocytes (Figure 2f), with a minor contribution of PD-L1 (Figure S2g). Of note, CD14^+^ARG1^+^ immune suppressive monocytes were originally defined in patients with pancreatic cancer who also share increased levels in some inflammatory cytokines [36, 37].

Further studies are necessary to define whether these cells are immature precursors of the granulocytic/monocytic lineage or a new subset of circulating monocytes, as well as the extent of their overlap with the immature CD14^+^ cells described in COVID-19 patients and sepsis [20, 33]. Noteworthy, our study identifies, new markers associated with disease severity in the peripheral blood of patients, such as the proportion of activated/resting γδ-T cells (Figure S3E), which integrate with others already described and confirmed in our study (i.e NK cells and neutrophils activation [38, 39]), which might be responsible for healthy tissue damage [40]. This information has crucial consequences from clinical and biological point of views and it will support physicians in patient stratification. More importantly, the extensive molecular analysis performed in BAL samples reveals that lymphoid compartment landscape perfectly mirrors compromised healthy conditions of the patients and identifies in the increase of naïve T cells in spite of CD4^+^ Trm and CD8^+^ Trm the worst clinical scenario (Figure 3C-D). Naïve T cells unbalance is not clearly observed in peripheral blood of the same patients suggesting that systemic analysis, despite many advantages, will not provide a complete landscape of the disease. According to this, we highlight the importance of CXCR6, which has has been recently associated with less severe COVID-19 stage [23]. CXCR6, highly expressed in Trm cells (Figure 3h), orchestrates Trm cells partitioning within the lung directing them to the airways [41].

Collectively, the deep molecular, phenotypic, and functional myeloid and lymphoid characterization performed on peripheral and local districts points out that critically ill patients face a profound immune dysregulated status, which can support secondary bacteria and virus infection (table 1, Figure 4c, Figure S4a). Indeed, as sign of immune paralysis, monocytes in septic patients do not respond to LPS stimulation with the upregulation of NF-κB-dependent genes, including TNFα [33]. Although this was not investigated in our study, we nonetheless show evidence that monocytes from patients who had a fatal outcome in ICU were dysfunctional and had lost the immune regulatory properties. It is thus likely that monocytes in terminally ill patients are flawed in different biological responses, possibly including the ability to differentiate into M1 macrophages in the lung, as observed in the BAL of these subjects. Together with the reset of lymphoid arm indicated by the relative abundance of naïve T cells, this configures a state of “immune silence” and supports the deploying of drugs that can “reawaken” host immune system. “Immune silence” could be related to the extensive immaturity of this cell population, as consequence of an abnormal and skewed myelopoiesis. Alternatively, it might be a pre-existing disorder making this group of patients unable to cope with the hyper-inflammatory state. Longitudinal studies are mandatory to dissect between these different disease developments.

Our data set the ground for the administration of drugs that aims at switching off and re-start host immune system in both adaptive and innate arms, as suggested by the efficacy of recently adopted dexamethasone in severe COVID-19 patients [42]. By combining scRNA-seq with functional assays, we were able to identify features associated with disease severity and clinical outcome of the patients. However, our study is not flawless and suffers from limitations, including experimental design ones. Firstly, this is not a longitudinal study. Secondly, the absence of BAL samples from mild patients prevents us to generalize and validate the observations made on severe ones. Moreover, the absence of BAL samples from healthy control deprives us of estimating the basal state and composition of the lung immune system. While it is technically possible to collect such samples, it raises important ethical question that need to be considered while designing COVID-19 cohorts. The third limiting factor is the nature of the BAL samples: briefly, a physiologic solution is introduced into the lower respiratory tract and then collected. Such method is able to capture infiltrating immune cells in spite of non-immune cell types (i.e. epithelial cells), as shown by recent autopsy reports [43]. Lung biopsy represents an interesting alternative to BAL but is far riskier, especially for patients suffering from ARDS, such as severe COVID-19 patients, and the amount of available material is limited. One may therefore try to collect samples from deceased patients, however as most patient died after several weeks in ICU, the biological features observed during the autopsy might either be linked to the disease itself or to the extended stay in ICU. In the case of lung biopsy, highly multiplexed imaging techniques such SeqFISH [44] or CODEX [45], but also spatial transcriptomic technologies [46] could be explored. Such approaches would provide gene expression spatial pattern at a near single-cell resolution and gain essential information concerning potential cellular interactions. Lastly, we observed that several patients had a significant interferon-induced polarization of blood neutrophils that could not be solely explained by the virus presence in the lung. Such polarizations might also be due to secondary infections, but mostly bacterial rather than viral. Therefore, an extensive metagenomic analysis of the BAL could reveal disease-associated bacterial landscape and be associated to specific immune phenotype.

Overall, our study expands the biological insight on the multifaceted virus-immune system interplay and provides a solid background to design and test new candidate drugs for severe COVID-19 patients.

## Methods

### Study subjects and clinical considerations

All 31 patients with COVID-19 and 5 healthy donors in this study were admitted, within the period from March 12th to April 20th 2020 to the University Hospital of Verona. At sampling, the stage of disease was categorized as mild (patients not requiring non- invasive/mechanical ventilation and/or admission to ICU) or severe (patients requiring admission to ICU and/or non-invasive/mechanical ventilation). This study includes a group of 21 severe COVID-19 patients admitted to ICU, 10 mild SARS-CoV-2 patients and 5 HDs. The clinical features of the 3 groups of subjects are recapitulated in table 1. The study has been designed with the purpose of defining a complete framework of COVID-19 patients’ immune landscape. To this aim we collected clinical (i.e. comorbidities, pulmonary performances, outcome at dismissal from Verona Hospital) and laboratory (i.e. leukocyte subsets, P-ferritin, P-D-Dimer, C-reactive protein, P-fibrinogen quantification) information and integrated them with molecular (i.e. single cell transcriptomic analysis) proteomic (cytokines quantification and serology), phenotypic (myeloid characterization in terms of expression of immune suppression hallmarks) functional (myeloid immune suppressive assay) data.

### Ethics approval statement

All the patients (and/or initially their families) provided written informed consent before sampling and for the use of their clinical and biological data. This study was approved by the local ethical committee (Prot. n° 17963, and n° 51095, P.I. Vincenzo Bronte); informed consent was obtained from all the participants to the study.

### Preparation of biological samples

For each patient approximately 20 ml of BAL fluid was obtained, stored at room temperature and processed within 2 hours in a BSL-3 laboratory. An unprocessed aliquot was used for bacterial culture. The BAL fluid was filtered 2 times through a nylon gauze and a 100-µm nylon cell strainer to remove clumps and debris. The supernatant was then washed with PBS 1x and centrifuged. RBCs were lysed with 4 mL of 0.2% NaCl solution (3 minutes, RT) and the reaction was blocked by adding 9 mL of 1.2% NaCl solution. The cells were washed with PBS 1x, re-suspended in RPMI 1640 medium supplemented with 5% bovine serum albumin and counted. Cell viability was determined by Trypan blue exclusion. BAL fluids of patients with COVID-19 infection contained a heterogeneous number of cells ranging from 0.83 ×10^6^ to 22 ×10^6^ of cells. Cells were re-suspended at a concentration of 1 × 10^6^ /ml for single cell analysis. Peripheral blood (PB) from COVID-19 patients and HDs was collected in EDTA-coated tubes. 2 ml of PB was washed once with PBS 1x and the RBCs lysis was performed twice adding 15 mL of 0.2% NaCl solution (3 minutes, RT) and the reaction was blocked by adding 35 mL of 1.2% NaCl solution. The cells were washed with PBS 1x, re-suspended in RPMI 1640 medium supplemented with 5% bovine serum albumin, filtered through a 100-µm nylon cell strainer and counted. Cell viability was determined by Trypan blue exclusion. Cells were resuspended at a concentration of 1 × 10^6^ /ml for single cell analysis.

### SARS-CoV-2 detection, bacteria identification and validation of HSV-1 infection

Nucleic acids were extracted from BAL samples by Seegene Nimbus instrument (Seegene; Seoul, South Korea) and SARS-CoV-2 related genes (E, N and RdRP) were amplified by polymerase chain reaction (PCR) with the Allplex 2019-nCoV assay kit (Seegene) according to the manufacturer’s instructions. BAL samples from COVID-19 positive patients were processed for bacterial isolation. Briefly, samples were treated with 1% v:v Dithiothreitol (DTT) for 30 minutes at 25 °C Upon treatment, (20 μl of) the samples were streaked out on petri dishes containing different types of agar (Blood Agar, Chocolate Agar, Columbia Nalidix Acid agar, Mannitol Salt Agar, McConkey and Sabouraud Agar). Cultures were incubated for 24 hours at 37 °C and bacterial growth was evaluated as colony forming unit per ml (CFU/ml). Bacterial species were identified by MALDI-tof (VITEK-MS, BioMérieux; France) and antimicrobial susceptibility was tested by VITEK-2, BioMérieux; France) and Kirby-Bauer disk diffusion assay. Nucleic acids isolated from BAL samples were processed to multiplex PCR procedure for the simultaneous detection of HSV-1, HSV-2, HCMV and VZV, using an Allplex assay (Seegene) according to the manufacturer’s instructions.

### Detection of cytokines and serology

Cytokines released by patients’ monocytes and neutrophils were quantified by Human Procarta-Plex™ Panel 1 multiplex (ThermoFisher Scientific, Waltham, MA, USA). The ELISA assay to detect Immunoglobulins (Ig) used fragment of the SARS-CoV2 spike glycoprotein (S-protein) as antigens based on a recently published protocol [47]. The Spike SARS-CoV2 glycoprotein receptor binding domain (RBD) was expressed in mammalian HEK293 cells at IEO, Milan by Drs. Marina Mapelli and Sebastiano Pasqualato as glycosylated proteins by transient transfection with pGACCS vectors generated in Dr. Krammer’s laboratory. The constructs were synthesized using the genomic sequence of the isolated virus, Wuhan-Hi-1 released in January 2020, and contained codons optimized for expression in mammalian cells. Secreted proteins were purified from the culture medium by affinity chromatography, quantified and stored in liquid nitrogen in aliquots. The ELISA tests to detect IgG in patients’ sera used as antigens the recombinant fragments of the RBD of the Spike SARS-CoV2 glycoprotein. After binding of the proteins to a Nunc Maxisorp ELISA plate, patients’ sera to be analyzed were applied to the plate to allow antibody binding, and then revealed with secondary antihuman- IgG (BD) antibody conjugated to HRP. Reaction was revealed upon addition of TMB (Merck). Optical density at 450 nanometers was measured on a Glomax (Promega) plate reader. All samples were tested and validated with an ELISA assay, as indicated in [48].

### Flow cytometry analysis

Immunophenotype analysis on whole peripheral blood was performed according to standard procedures in order to characterize monocyte subsets (defined as classical, CD14^high^ CD16^low/dim^; intermediate CD14^int^ CD16^+^; non classical CD14^low/dim^ CD16^high^). Briefly, peripheral blood was incubated with FcR Blocking reagent (Miltenyi Biotec, Paris, FR) followed by the addition of: antihuman PE-conjugated CD56 (BD Bioscience, San Jose, CA, USA; clone NCAM16.2), FITC-conjugated-CD16 (Biolegend, San Diego, CA, USA; clone 3G8), PerCP-Cy5.5-conjugated CD3 (BD Bioscience, San Jose, CA, USA; clone UCHT1), PE.Cy7-conjugated HLA-DR (eBiosciences, ThermoFisher Scientific, Waltham, MA, USA; clone L243), APC.H7-conjugated CD14 (BD Bioscience, San Jose, CA, USA; clone MφP9), Brilliant Violet 421™-conjugated PD-L1 (BD Bioscience, San Jose, CA, USA; clone MIH1) and aleza fluor 647-conjugated ARG1 (developed in our laboratory [36]) antibodies, Aqua LIVE/DEAD dye (ThermoFisher Scientific, Waltham, MA, USA). RBCs were lysed using Cal-Lyse™ Lysing Solution (ThermoFisher Scientific, Waltham, MA, USA) in accordance with the manufacturer’s instructions. Samples were acquired with FACS Canto II (BD, Franklin Lakes, NJ, USA) and analyzed with FlowJo software (Tree Star, Inc., Ashland, OR, USA).

### Myeloid cell isolation, phenotypic characterization and functional assay

Cells were isolated from EDTA-treated tubes (BD Biosciences, NJ, USA) and freshly separated by Ficoll-Hypaque (GE Healthcare, Uppsala, Sweden) gradient centrifugation. PBMCs were counted and the monocyte fraction (CD14^+^) was further isolated by CD14- microbeads (Miltenyi), following manufacturer’s instructions. From the CD14- fraction the CD66^+^ low density gradient neutrophils (LDNs) were isolated by the sequential addition of CD66b-FITC antibody (BD Biosciences, NJ, USA) and microbeads anti-FITC (Miltenyi), following manufacturer’s instructions. The normal density neutrophils (NDNs) CD66b^+^ were isolated from the RBC layer by dextran density gradient followed by CD66b-antibody as described for LDNs. The purity of each fraction was evaluated by flow cytometry analysis. Samples with a purity greater than 95% were assessed for their suppressive capacity. 0.5×10^6^ cells of each cell type were plated in 24-well plates for 12 hours in complete RPMI supplemented with 10% FBS. At the end of the incubation, viability was evaluated by flow cytometry and Trypan blue assay, and both the supernatants and the cells were collected and cultured with CellTrace (Thermo Fisher Scientific) labeled PBMCs, stimulated with coated anti-CD3 (clone OKT-3, eBioscience, Thermo Fisher Scientific) and soluble anti-CD28 (clone28.2, eBioscience, Thermo Fisher Scientific) for 4 days in 37°C and 8% CO2 incubator (for details refer to [49]. For the cells a ratio of 3:1 (target:effector) was used. At the end of the culture, cells were stained with anti-CD3-PE/Cy7 (UCHT1, eBioscience, Thermo Fisher Scientific) and CellTrace signal of lymphocytes was analyzed with FlowJo software (Tree Star, Inc. Ashland).

### Single-cell RNA-sequencing (scRNAseq)

BAL and peripheral blood cells were isolated and prepared as described above. For each sample, cells were resuspended in RPMI supplemented with 5% FBS to a final concentration of 1000 cells per ml and processed using the 10x Genomics Chromium Controller and the Chromium NextGEM Single Cell 3′ GEM, Library & Gel Bead kit v3.1 (Pleasanton, California, United States) following the standard manufacturer’s instructions. In brief, 10,000 live cells were loaded onto the Chromium controller to recover 4,000 single cell GEMs per inlet uniquely barcoded. After synthesis of cDNA, sequencing libraries were generated. Final 10X library quality was assessed using the Fragment Analyzer High Sensitivity NGS kit (Agilent Technologies, Santa Clara, CA, USA) and then sequenced on the Illumina NextSeq500 (Illumina, San Diego CA, USA) generating 75 base pair paired-end reads (28bp read1 and 91bp read2) at a depth of 50,000 reads/cell.

### Data analysis and statistics

#### Generation of UMI tables

Upstream processing of reads was done using the CellRanger toolkit with default parameters. SARS-CoV-2 (NCBI reference number: NC_045512.2) and human hg38 genomes were downloaded from NCBI website. SARS-CoV-2 GTF annotation file was downloaded from the UCSC and merged with the human GTF as an additional chromosome. ORF_10 Gene 3’ boundary extended by 100 bases to catch all reads that belong to this transcript.

#### High-level analysis of scRNA-seq expression data

ScRNA-seq expression data analysis were performed using the R-based Pagoda2 pipeline (https://github.com/hms-dbmi/pagoda2/) (29227469) in addition to an in-house R script. Briefly UMI table were loaded using the read.10x.matrices() function. Low quality cells were removed using the following strategy: cell with less than 500 UMIs and more than 20% of mitochondrial genes were removed. Two rounds of analysis were performed: in the first one, all filtered cells were used to identify the major cell types, then cells from each cellular compartments are analyzed individually to provide more detailed informations. For each analysis, the number of Highly Variables Genes (HVGs) was determined using the adjustVariance() function with the gam parameter set to 10. HVGs were selected using the following strategy: for each gene, its number of zeros and its mean expression are computed. A local polynomial model is then used to predict the number of zeros according to the log mean expression (loess function with degree parameter set to 2). The residuals of this model (excess of zeros) are then used to ranked the genes and the genes with the highest excess of the zeros are considered as the most HVGs. PCA reduction is then computed using the calculatePcaReduction() function. The number of computed PC was changed in each analysis due to variable number of cells and cellular heterogeneity. A K-nearest neighbor graph was then build with the function makeKnnGraph() with the K value set to 30 and the distance parameter set to ‘cosine’. In order to get high-quality cell clusters, we used the Leiden community detection implemented in the R package **leiden**, a wrapper of the python package **leidenalg**. The leiden() function was applied to the KNN graphs with default parameters for each analysis. Marker genes were identified using the getDifferentialGenes() function. UMAP low dimensional embedding was computed using the **uwot** R package, and more precisely the umap() function with the n_neighbors parameter set to 30, and the metric parameter set to ‘cosine’. In order to group clusters of cells in the first round of analysis, mean gene expression of the most variable genes was computed using the aggregate() function. Spearman’s correlation matrix was then computed using the cor() function with the method parameter set to ‘Spearman’. Hierarchical clustering was then performed on this matrix using Ward’s method and the resulting tree used to aggregate the cell clusters.

#### Correspondence Analysis of the scRNA-seq data

In order to identify trends in cellular composition across samples we used a multivariate technique called Correspondance Analysis (CA). CA is conceptually similar to principal component analysis but applies to categorical rather than continuous data. It is traditionally applied to contingency tables: CA decomposes the chi-squared statistic associated with this table into orthogonal factors. Because CA is a descriptive technique, it has the advantage of being applicable to tables whether or not the chi-squared statistic is appropriate. We used the R implementation of CA from the package **FactoMineR** (CA function) with default parameters. To determine the significant components we looked at the scree plot and selected the eigenvalues/component located before the elbow. To improve the quality of our analysis, we removed cell clusters corresponding to red blood cells, platelets and cancer cells from patient 8.

To detect clinical and biological variables associated with the computed correspondance components we used the following strategy: for cytokine concentrations, we first took the square root of the initial values to get normally distributed variables and then computed Pearson’s correlation with each component independently. For the other continuous variables (clinical scores, age, BMI…), Pearson’s correlation was directly computed. To test the association between CA component or a specific cell type proportion and a categorical variable (i.e. patient clinical status and survival) we either applied a Tukey’s range test (TukeyHSD() function) if the variable is not heavy tailed. If the cell proportions are clearly heavy-tailed, we applied a Kruskall-Wallis rank test.

#### Viral-Track analysis

To detect and study viruses in our scRNA-seq samples we used Viral-Track, a computational tool that screen the raw sequencing files to find viral reads (32479746). As previously described, processing of the file was performed using UMI-tool (28100584). First, cell barcodes were extracted and a putative whitelist computed using the umi_tools whitelist command with the parameters ‘–stdin —bc-pattern = CCCCCCCCCCCCCCCCNNNNNNNNNN –log2stderr’. Following the mapping of the reads to viral genomes and transcript assembly, the mapped reads were assigned to transcripts using the R package Rsubread through the function featureCounts() with default parameters. The command ‘umi_tools count’ is then used to compute the final expression table with the following parameters:–per-gene–gene-tag = XT–assigned-status-tag = XS–per-cell.

In the case of patient 8, cells were not filtered on total host UMIs and proportion of MT UMIs but only on total combined host and viral UMIs to avoid removing apoptotic cells containing a high viral load but expressing few host genes.

#### Analysis of the serum cytokine, blood cell count and clinical data

Using a Cullen and Frey graph (descdist() function from the **fitdistrplus** package)we observed that both serum cytokine and blood cell count variables could be transformed into gaussian-like variables by applying a simple square root function and then used for further analysis. Association between blood cell counts or serum cytokine concentration and patient clinical status was assessed by fitting an ANOVA model to the transformed variables (aov() and anova() functions). Correction for multiple testing was done using the p.adjust function with parameter method set to ‘BH’. When correlations with a CA dimension were computed, the cor() function with default parameters was used. To validate the association between the SOFA score and the lymphoid CA dimension 1 we fitted a basic linear model with the lm() function and assessed the significance of the association by performing a Fisher test with the anova() function.

#### Analysis of the immuno-suppression, flow cytometry and cytokine secretion data

As both both flow cytometry and cytokine secretion data were extremely heavy-tailed we applied a logarithmic transformation with a pseudo count of 1 (log10(1+x)). Spearman correlations between protein MFI or cytokine concentration and immune-suppression was computed using the cor() function.

In order to model the relationship between ARG1 MFI and immune-suppression we used a function similar to the Hill function used in biochemistry:

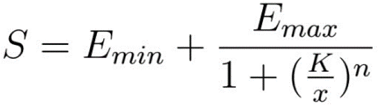

Here *S* corresponds to the immune-suppression, x to the transformed ARG1 MFI, *E_min_* to the basal immune-suppression, E_max_ to the maximal suppression that can be induced by ARG1, *K* to the transformed ARG1 MFI required to get half of the maximal suppression (*E_min_*+*E_max_*) and *n* the cooperation coefficient. This function was fitted using the nls() function with default parameters.

Quantitative variables indicated in table 1 were expressed as the median and interquartile range (IQR), qualitative ones as percentages. All statistical analysis were performed using R 3.6.1 on an Ubuntu 18.04 workstation.

## Reporting summary

Further information on research design is available in the Nature Research Reporting Summary linked to this paper.

## Data availability

The authors declare that all the other data supporting the findings of this study are available within the article and its supplementary information files and from the corresponding authors upon reasonable request. All scripts used for the data analysis are available at https://github.com/PierreBSC/Verona_COVID19.

## Data Availability

The authors declare that all the other data supporting the findings of this study are available within the article and from the corresponding authors upon reasonable request.

## Acknowledgement

This work was supported by Fondazione Cariverona (ENACT Project) and Fondazione TIM.

We thank all patients who participated in this study and their families. We thank all the members of Immunology Section of Verona University Hospital who actively worked during the pandemia: Francesca Hofer, Varvara Petrova, Chiara Musiu, Cristina Frusteri, Veronica Batani, Alessandro Ghirardelli, Morena Martini, Fiorenza Paiola, Elena Lucchini, Claudia Pizzoli, Elena Chiesa, Oretta Gabrielli, Nadia Brutti, Monica Brentegani, Elisabetta Gallo, Giulio Fracasso, Tiziana Cestari, Ornella Poffe, Antonio Vella, Giovanna Zanoni, Silvia Sartoris, Riccardo Ortolani, Selena Gomirato, Daniel Lovato, Antonella Valentini and Claudia Italia. We thank the members of Division of Infectious Diseases who actively assist patients: Monica Brentegani, Alessandro Visentin, Amina Zaffagnini and Leonardo Motta. We thank the members of Intensive Care Unit who were actively involved in patients care and assistance: Sara Boschetti, Francesca Del Favero, Riccardo Boetti. We thanks Marina Mapelli and Sebastiano Pasqualato of European Institute of Oncology for technical support. We thank 10X Genomics Inc. and Carlo Erba to support the research. We deeply acknowledge the contribution of ‘Centro Piattaforme Tecnologiche” of the University of Verona for samples sequencing.

We dedicate this work to the memory of health care workers who have given their lives in the care of patients with COVID-19.

## Author information

These authors contributed equally: Pierre Bost, Francesco De Sanctis and Stefania Canè

These author share senior contribution: Ido Amit and Vincenzo Bronte

### Author Contribution

P.B. designed and developed Viral-Track, performed computational and statistical analysis and wrote the manuscript. F.D.S. performed sample processing, bio banking and single cell experiments, coordinated the study, data analysis and wrote the manuscript. S.Can. performed sample processing, bio banking, functional studies and data analysis. S.U. coordinated the study and wrote the manuscript; K.D. was responsible for biological specimens and patients’ data collection and revised the manuscript; M.C. performed single cell experiments; D.E. contributed to data processing and analysis; A.F., C.A., R.M.B. and R.T. performed sample processing, bio banking, functional studies and data analysis; S.Cal contributed to computational analysis; A.L., M.I. and F.F. performed cytokine and serological analysis and revised the manuscript; D.G., A.R.M., were responsible for microbiological analysis and revised the manuscript; P.D.N., E.T., L.G. and E.P. were responsible for biological specimens and patients’ data collection and revised the manuscript; B.S. contributed to development of computational methods and bioinformatic analysis and revised the manuscript; I.A. and V.B. directed the project, conceived, designed the experiments, and wrote the manuscript.

### Corresponding Authors

Ido Amit

Vincenzo Bronte

